# Comparative Efficacy and Safety of Tenecteplase and Alteplase in Acute Ischemic Stroke: A Pairwise and Network Meta-analysis of Randomized Controlled Trials

**DOI:** 10.1101/2022.10.06.22280794

**Authors:** Aqeeb Ur Rehman, Aleenah Mohsin, Huzaifa Ahmad Cheema, Afra Zahid, Muhammad Ebaad Ur Rehman, Muhammad Zain Ameer, Muhammad Ayyan, Muhammad Ehsan, Abia Shahid, Muhammad Aemaz Ur Rehman, Jaffer Shah, Ayaz Khawaja

## Abstract

**Background:** Studies on tenecteplase have been yielding mixed results on several important variables at different doses, thus hampering objective guideline recommendations in acute ischemic stroke management. This meta-analysis stratifies doses in order to refine our interpretation of outcomes and quantify the benefits and harms of tenecteplase at different doses.

**Methods:** PubMed/MEDLINE, the Cochrane Library, and reference lists of the included articles were systematically searched. Several efficacy and safety outcomes were pooled and reported as risk ratios (RRs) with 95% confidence intervals (CIs). Network meta-analysis was used to find the optimal dose of tenecteplase. Meta-regression was run to investigate the impact of baseline NIHSS scores on functional outcomes and mortality.

**Results:** Ten randomized controlled trials with a total of 4140 patients were included. 2166 (52.32%) patients were enrolled in the tenecteplase group and 1974 (47.68%) in the alteplase group. Tenecteplase at 0.25 mg/kg dose demonstrated significant improvement in excellent functional outcome at 3 months (RR 1.14, 95% CI 1.04-1.26), and early neurological improvement (RR 1.53, 95% CI 1.03-2.26). There was no statistically significant difference between tenecteplase and alteplase in terms of good functional outcome, intracerebral hemorrhage (ICH), symptomatic intracerebral hemorrhage (sICH), and 90-day mortality at any dose. Meta-regression demonstrated superior tenecteplase efficacy with increasing stroke severity, however, the results were statistically nonsignificant.

**Conclusions:** Tenecteplase at 0.25 mg/kg dose is more efficacious and at least as safe as alteplase for stroke thrombolysis. Newer analyses need to focus on direct comparison of tenecteplase doses and whether tenecteplase is efficacious at longer needle times.

## Introduction

Stroke is a leading cause of morbidity, mortality, and long-term disability globally (1). The currently approved approach to the management of acute ischemic stroke consists of thrombolytic therapy with alteplase, an established first-line therapy at a dose of 0.9 mg/kg, 10% of which is administered as a bolus, and the rest is given as a continuous infusion over an hour (2). Recently, tenecteplase, a modified variant of alteplase, has emerged as a promising alternative to alteplase due to a longer half-life (slower plasma clearance), greater specificity for fibrin, and reduced binding affinity to Plasminogen Activator Inhibitor (PAI) (3,4). Furthermore, its administration as a single bolus makes it a more convenient alternative to alteplase for both hospital staff and patients, and allows administration in mobile stroke units, thus reducing needle time and improving outcomes (5). These findings call for a reevaluation of alteplase as the first-line therapy for acute ischemic stroke.

Randomized controlled trials (RCTs) exploring comparative efficacy and safety of tenecteplase and alteplase have been yielding contradictory and inconsistent results on various outcomes and merit the conduction of a meta-analysis to adequately answer these questions which may have huge real-life impact on stroke outcomes. The most recent meta-analysis published earlier in 2022 is limited by smaller sample size and inclusion of observational studies which add to the unreliability of results owing to their inherent vulnerability to information bias and confounding (6). In addition, the publication of AcT trial adds the single largest pool of data in comparing tenecteplase and alteplase (7), necessitating a systematic compilation of all available RCTs on the topic, potentially with greater confidence in results. The previous meta-analysis also fails to stratify analysis according to doses of tenecteplase and categorizes all three doses (0.1 mg/kg, 0.25 mg/kg, 0.4 mg/kg) as identical under one tier in reporting their results, further amplifying the need for a newer meta-analysis that can differentiate between different doses of tenecteplase in addition to comparing tenecteplase to alteplase.

## Methods

This meta-analysis conforms to the guidelines elucidated in Preferred Reporting Items for Systematic Reviews and Meta-Analyses (PRISMA) statement (8) and has been registered with the International Prospective Register of Systematic Reviews PROSPERO (CRD42022355362).

### Literature Search

We used MEDLINE (via PubMed), the Cochrane Controlled Register of Trials (CENTRAL) (via the Cochrane Library), and clinicaltrials.gov up till September 2022 to search for studies that compared tenecteplase with alteplase in stroke patients. The detailed search strategy is available in the Supplementary File. No restriction in terms of language, time, or location was applied while searching the literature. Additionally, reference lists of previous meta-analyses and included trials were manually screened to identify any further relevant studies.

### Study selection and data extraction

Randomized controlled trials (RCTs) that compared tenecteplase with alteplase in adult patients (age ≥18) undergoing thrombolysis for stroke were selected for this meta-analysis. All records were imported to Mendeley Desktop where two authors working independently screened them, first based on titles and abstracts, and then by reading full texts for the remaining articles. Any disagreements were resolved by a third author. Two authors independently applied a predesigned data collection sheet to extract the following: demographic variables such as age and sex, length of hospital stay, median NIHSS at baseline, needle time, intervention (tenecteplase), tenecteplase dose, and various efficacy and safety outcomes at those doses. Efficacy outcomes included functional status at three months (mRS 0-1 and mRS 0-2), early neurological improvement, and recanalization. The safety outcomes of interest were intracranial hemorrhage (ICH), symptomatic intracranial hemorrhage (sICH), and mortality at 3 months.

### Risk of bias assessment in individual studies

The risk of bias in the included studies was assessed using the revised Cochrane “Risk of bias” tool for randomized controlled trials (RoB 2.0) (9). Articles were assessed across five domains: (1) bias arising from the randomization process; (2) bias due to deviations from intended interventions; (3) bias due to missing outcome data; (4) bias in the measurement of the outcome and (5) bias in the selection of the reported result. Two review authors independently assessed the risk of bias for each study. In the case of any disagreements, a third review author was consulted.

### Statistical analyses

We reported dichotomous outcomes as relative risk (RR) at 95% confidence interval. The meta-analysis was performed using DerSimonian and Laird random-effects model. We performed subgroup analyses based on the dose of Tenecteplase (0.1 mg, 0.2-0.25 mg, and 0.32-0.4 mg). For each analysis, we calculated the Chi^**2**^ and I^**2**^ statistics to assess the presence of heterogeneity and quantify it, respectively. The statistical analysis was performed using Review Manager (RevMan, Version 5.4).

To explore the causes of heterogeneity, a leave-one-out sensitivity analysis and meta-regression were performed. We performed meta-regression for functional outcomes and mortality using OpenMetaAnalyst software with the baseline NIHSS scores as covariates.

Additionally, we performed a network meta-analysis on our primary outcomes to assess the optimal dose of tenecteplase with the alteplase group as comparator using MetaXL 5.3. MetaXL implements Generalized Pairwise Modeling (GPM) framework for network meta-analysis. This framework is based on the Bucher method which involves repeated application of adjusted indirect comparisons (10,11).

## Results

### Study selection

A comprehensive database search yielded a total of 892 records, while 1 article was identified through additional sources. After removing duplicates (n=157), the remaining articles were subjected to primary screening based on titles and abstracts resulting in the exclusion of a further 708 studies. The full texts of remaining articles were screened in accordance with the inclusion criteria laid out in the study protocol, eventually yielding 10 RCTs for inclusion in the quantitative synthesis. The PRISMA flowchart is summarized in Figure 1.

**Figure 1.**
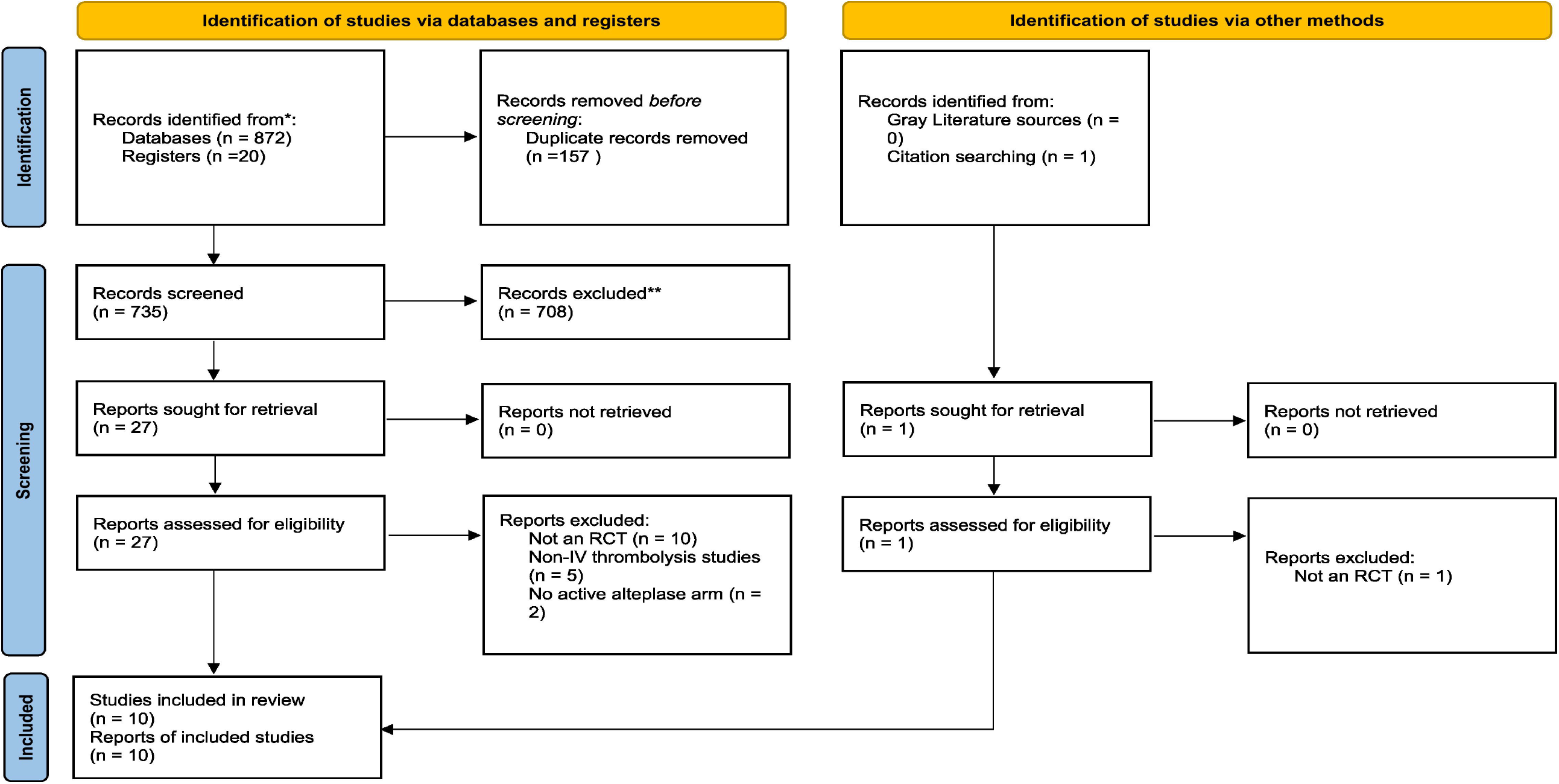
PRISMA 2020 flowchart.

### Study and Patient characteristics

After an extensive literature review, we included 10 studies spanning from 2010 to 2022 (7,12–19). 4140 patients were included, of whom 2166 (52.31%) were enrolled in the tenecteplase group and 1974 (47.68%) in the alteplase group. Across all included studies, we found 3 cohorts that tested for tenecteplase efficacy and safety at a dosage of 0.1 mg/kg, 8 cohorts that tested for TNK at dosages of 0.2-0.25 mg/kg, and 4 cohorts that tested for dosages of 0.32-0.4 mg/kg. Across all trials, the dosage of alteplase received by all control group participants was 0.9 mg/kg.

Mean needle time i.e. the time between stroke onset and thrombolysis, varied considerably among included trials but a majority of the trials defined cut-off at <3 hour. Exceptions to this included Bivrad et al., Kvistad et al., and Huang et al. who used a time window of <4.5 hours. The baseline National Institutes of Health Stroke Scale (NIHSS) also varied among the studies, however with the exception of Logallo 2017 and Campbell 2018, all fell in the moderate stroke severity range of the scale (5-15). Table 1 summarizes the characteristics of included studies.

**Table 1:**
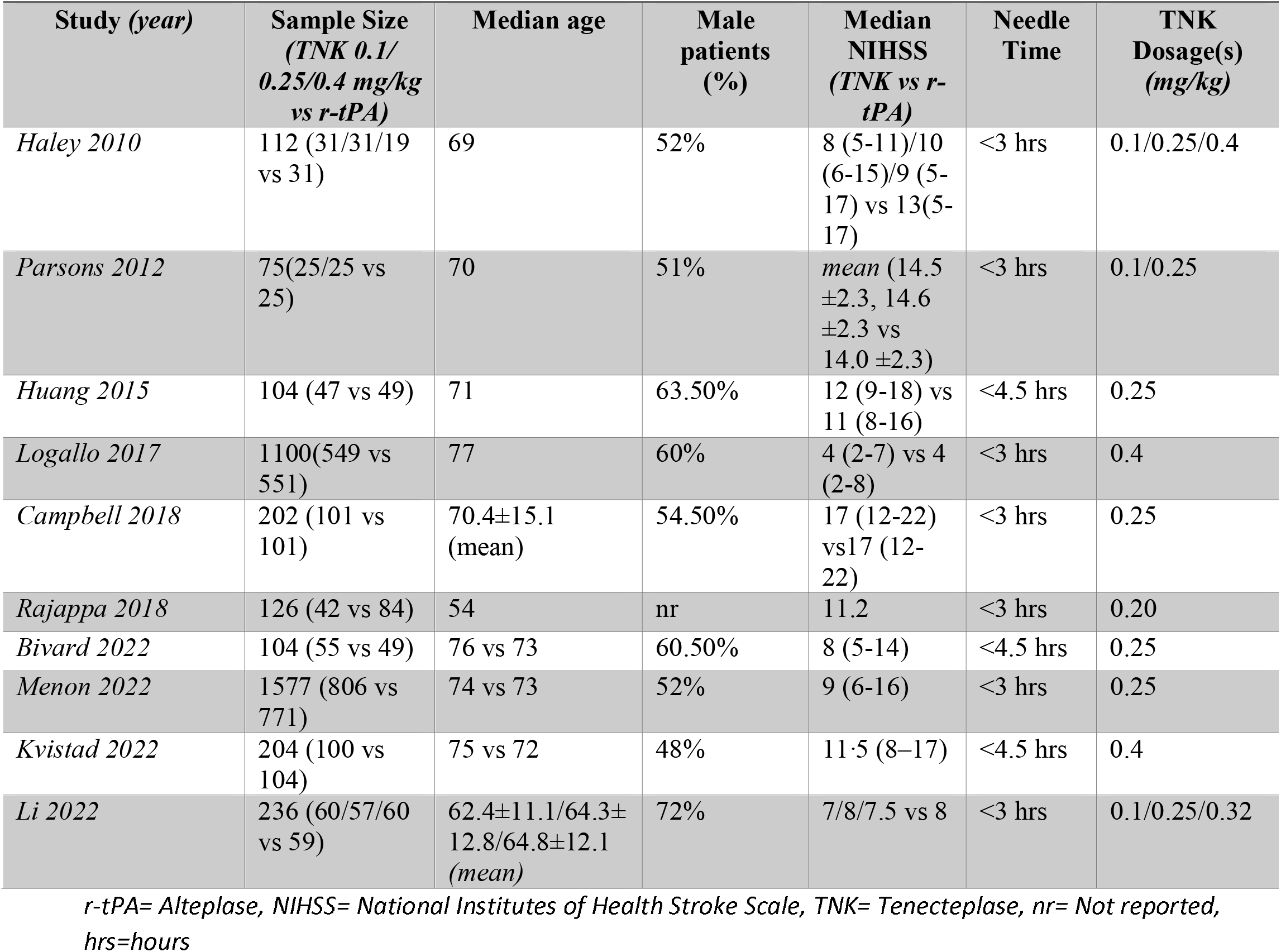
Characteristics of Included Studies.

### Quality Assessment of Included Studies

Overall, three trials were judged to have some concerns of bias due to issues in the domains of randomization process and selection of the reported result (Figure 2). The rest of the trials were found to be at a low risk of bias in all domains.

**Figure 2.**
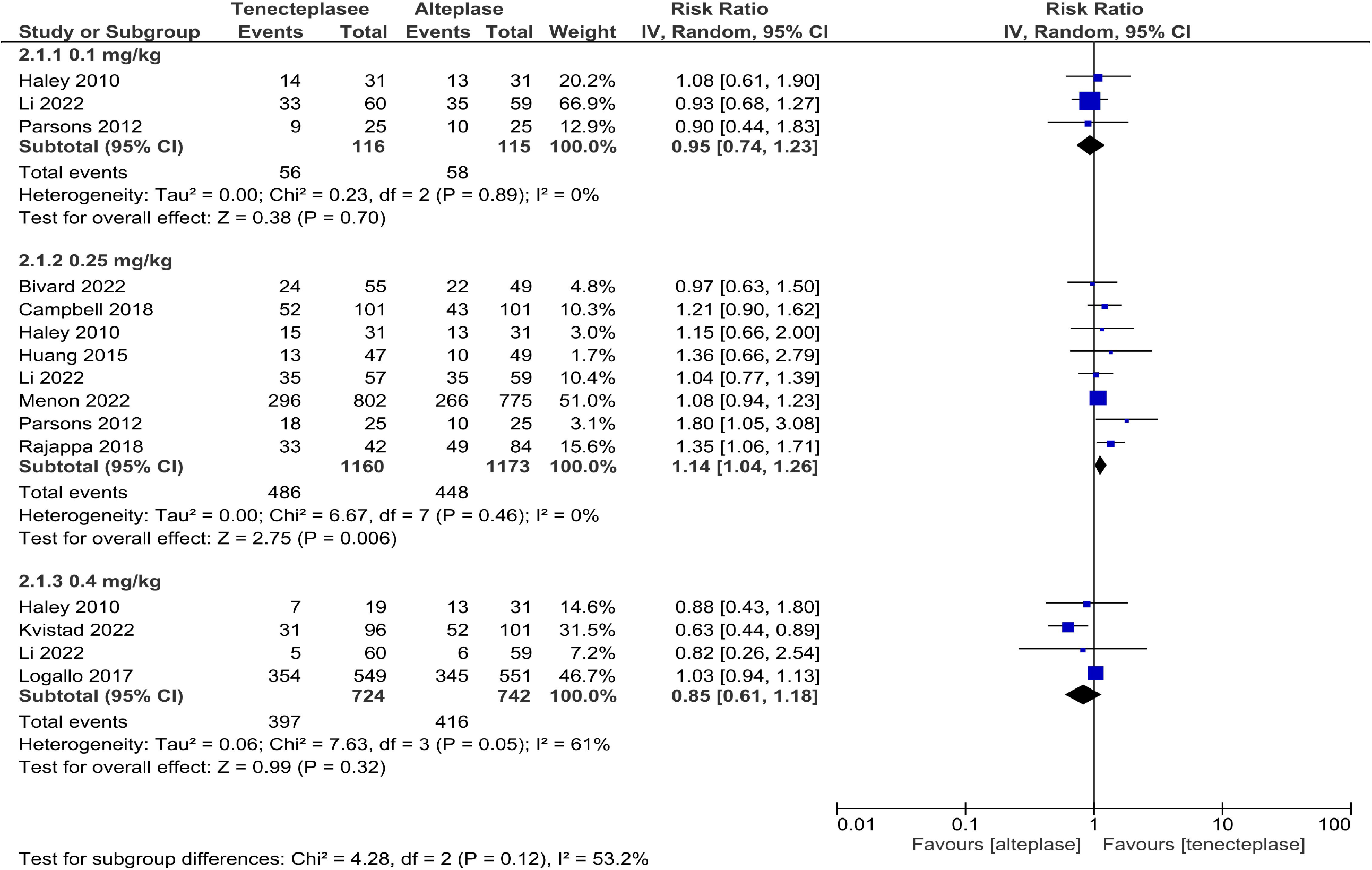
Quality assessment of included RCTs.

### Efficacy outcomes

#### 1. Functional outcomes

Significant improvement was observed for excellent functional outcome at 3 months (mRS 0-1) between the experimental and control groups (RR 1.14, 95% CI 1.04-1.26, I2=0%) when data were analyzed from 8 different trials encompassing 2333 patients in the 0.25 mg/kg groups. However, the rates of 90-day excellent outcome did not differ statistically in the groups receiving tenecteplase at 0.1 and 0.4 mg/kg doses (RR 0.95, 95% CI 0.74-1.23, I^2^ =0%; RR 0.85, 95% CI 0.61-1.18, I^2^ =61% respectively; Figure 3).

**Figure 3.**
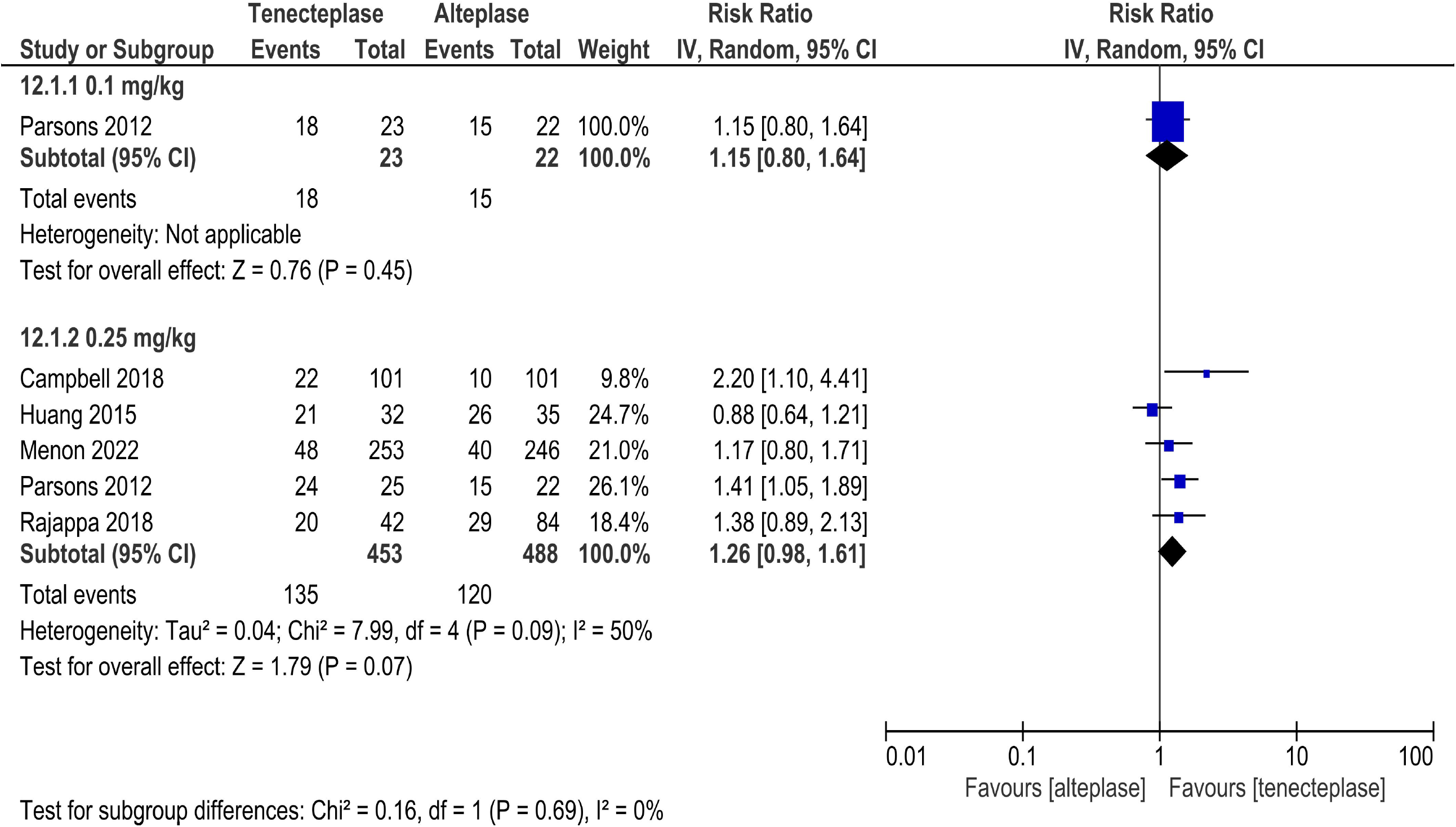
Forest plot comparing tenecteplase with alteplase in terms of excellent functional outcome.

Sensitivity analysis in the 0.4 mg/kg cohort upon removing the NOR-TEST trial from the analysis removed the heterogeneity with the pooled estimate favoring alteplase significantly (RR 0.68, 95% CI 0.50-0.92, I^2^ =0%).

Considering all three dose tiers of tenecteplase group, results for good functional outcome (mRS 0-2 at 3 months) came out to be nonsignificant after collating data from 8 trials that reported mRS 0-2, suggesting no improvement in good functional outcome at any dose. The summary RR for each of the three tiers (0.1mg/kg, 0.2-0.25mg/kg, 0.32-0.4mg/kg) were 1.04 (95% CI 0.75-1.44, I2 =35%), 1.11 (95% CI 0.98-1.26, I^2^ =47%), 0.86 (95% CI 0.69-1.08, I^2^ =79%) respectively, indicating non-inferiority of tenecteplase to alteplase at all doses (Supplementary Figure 1).

Meta-regression by baseline NIHSS scores for all doses combined demonstrated superior tenecteplase efficacy with increasing stroke severity, however, the results were nonsignificant for both excellent and good functional outcomes (p-value=0.258; p-value=0.271, respectively; Supplementary Figures 2 and 3). Meta-regression was also run for the 0.25mg/kg dose tier separately, yielding a stronger trend but nevertheless, nonsignificant results (p-value=0.171; p-value=0.084, respectively; Supplementary Figures 4 and 5).

#### 2. Early neurological improvement

Significant neurological improvement was demonstrated only by patients who were administered tenecteplase at the dose of 0.25 mg/kg (RR 1.53, CI 1.03-2.26, I^2^ =64%). Nonsignificant results were observed in the other two tiers, with 0.1 mg/kg cohort trending in favor of tenecteplase and 0.4 mg/kg cohort trending in favor of alteplase (Figure 4).

**Figure 4.**
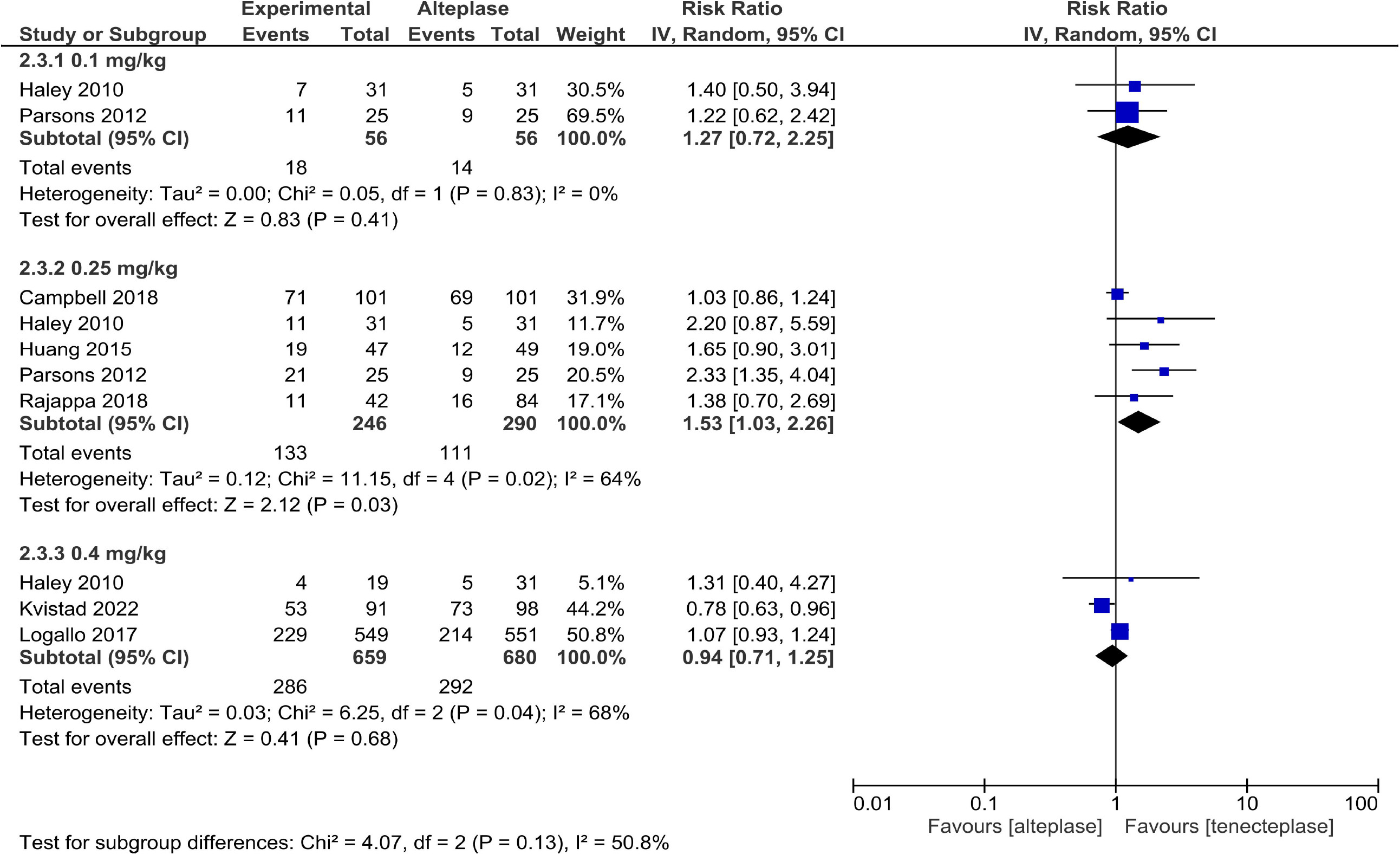
Forest plot comparing tenecteplase with alteplase in terms of early neurological improvement.

Heterogeneity in the 0.4 mg/kg group (I^2^=68%) was removed by the exclusion of the NOR-TEST trial, significantly altering the results in favor of alteplase.

#### 3. Recanalization

Limited data on recanalization hindered the attempt to run analyses on 0.4 mg/kg tier. Nonsignificant results were obtained for both 0.1 mg/kg group and 0.25 mg/kg group (RR 1.15, 95% CI 0.80-1.64; RR 1.26, 95% CI 0.98-1.61, I2 =50% respectively) (Figure 5).

**Figure 5.**
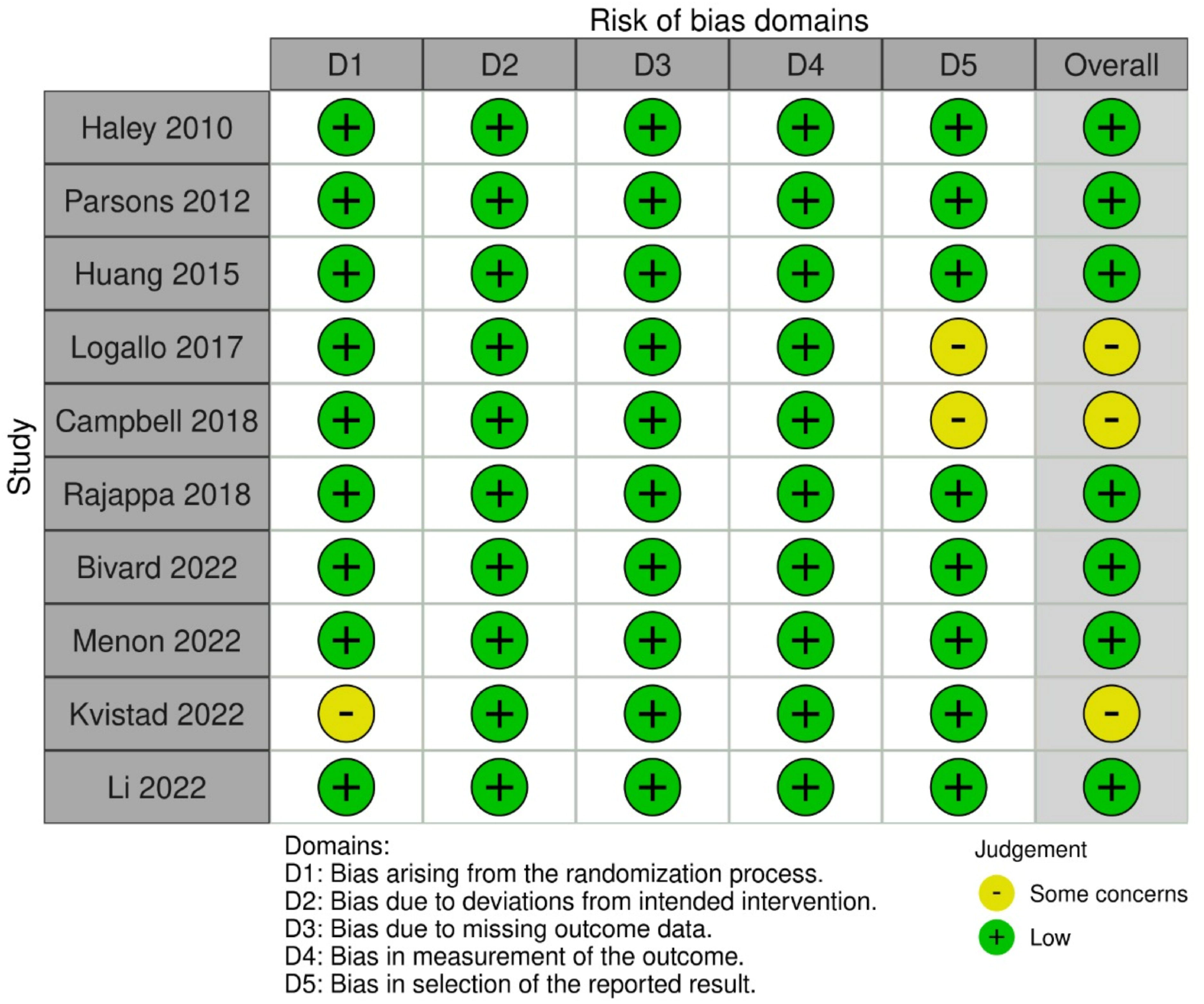
Forest plot comparing tenecteplase with alteplase in terms of recanalization.

### Safety outcomes

All-cause mortality rates did not differ statistically at any dose of tenecteplase when compared with alteplase (Supplementary Figure 6). However, there was a trend towards benefit with tenecteplase for the 0.1 mg/kg and 0.25 mg/kg doses. Meta-regression by baseline NIHSS scores, for all doses combined and separately for 0.25mg/kg dose tier, yielded nonsignificant results (p-value=0.356 for all doses combined; p-value=0.264 for 0.25mg/kg dose tier; Supplementary Figures 7 and 8).

Intracerebral hemorrhage (ICH) and Symptomatic Intracerebral Hemorrhage (sICH) when compared between the two groups, did not reveal statistically significant results, demonstrating noninferiority of tenecteplase (Supplementary Figures 9 and 10). However, the results in the 0.4 mg/kg tier trended in favor of alteplase.

### Results of the network meta-analysis

The results of our network meta-analysis showed that the 0.25 mg/kg dose of tenecteplase was associated with the greatest improvement in the functional outcomes although the results were statistically nonsignificant (Supplementary Figures 11 and 12). There was a trend towards an increased risk of sICH with the 0.4 mg/kg dose while the other dose tiers demonstrated a nonsignificant reduction in the risk of sICH as compared to alteplase (Supplementary Figure 13). The results for mortality were similar for all doses (Supplementary Figure 14).

## Discussion

A superior pharmacokinetic and pharmacodynamic profile of tenecteplase when compared with alteplase warrants serious consideration of tenecteplase as an alternative to alteplase. Our results show that the efficacy of tenecteplase exceeds alteplase in terms of several important outcomes; even with outcomes that are non-significant, there is a trend towards benefit with the administration of tenecteplase. Nevertheless, the statistical significance and strength of these trends notably differ at different doses of tenecteplase, further reinforcing the predetermined rationale of our study.

To the best of our knowledge, this is the largest meta-analysis to date that evaluates the efficacy and safety of different doses of tenecteplase for acute ischemic stroke. We found statistically significant improvement in excellent functional outcome at 3 months and early neurological improvement only with the 0.2-0.25 mg/kg dose tier of tenecteplase, indicating its superiority over other doses. Contrary to previous meta-analyses that included observational studies to find similar results for early neurological improvement (6,20), we have greater confidence in our results owing to the inherent strength of evidence of RCTs and an absence of heterogeneity with the 0.2-0.25 mg/kg dose tier. However, there was significant heterogeneity in the 0.4 mg/kg dose subgroup (I^2^>60% for excellent functional outcome; I^2^>75% for early neurological improvement) which disappeared upon the exclusion of NOR-TEST. This heterogeneity was likely the result of a lower baseline median NIHSS score of 4 in NOR-TEST, indicating a population of predominantly milder stroke patients compared to the other RCTs that recruited patients only with moderate or severe stroke (NIHSS 5-15; 16-20).

In contrast to alteplase which is known to have better efficacy in patients with mild stroke (21,22), results from our meta-regression showed that there was a statistically nonsignificant trend towards increasing benefit with tenecteplase in both excellent and good functional outcomes with increasing stroke severity. This is consistent with Kheiri et al. who showed that there was a significant improvement in the poor recovery outcome (mRS 4-6) as the baseline median NIHSS increased (23). However, given the statistical nonsignificance of this trend in our study, it is imperative to interpret it with caution and explore this association further in upcoming meta-analyses.

The American Stroke Association (ASA) guidelines recommend IV thrombolysis in all eligible patients irrespective of whether they are planned for undergoing thrombectomy (24). The results of pooled analyses have been inconsistent in the past, with some studies finding bridging therapy superior to thrombectomy alone (25). However, more recent meta-analyses now demonstrate similar efficacy and worse safety outcomes with bridging therapy, thus favoring thrombectomy alone as a substitute to thrombolysis (26). This is especially important given the obvious benefits of thrombectomy in terms of faster restoration of blood flow, shorter hospital stay, lower risk of intracerebral hemorrhage, and no contraindication in terms of age (27). In light of the ongoing debate on whether thrombolysis is even relevant today, it is important to point out that the trials investigating the subject have all predominantly employed alteplase and not the potentially more beneficial tenecteplase (28). Notably, contrary to previous analyses (6,20,23), our study did not find any difference in recanalization as well as safety outcomes between alteplase and tenecteplase at any dose; this could potentially mean that thrombectomy is likely superior to tenecteplase as well. However, our results are limited by a scarcity of data in this domain, and hence must not be generalized until a direct comparison of tenecteplase with thrombectomy is performed in clinical trials.

All safety outcomes including ICH, sICH, and all-cause mortality at 3 months came out to be nonsignificant at all doses in both the pairwise analysis and network analysis, corroborating the piling evidence in favor of noninferiority of tenecteplase to alteplase. The NOR-TEST 2 trial found an increased risk of cerebral hemorrhage and 90-day mortality at 0.4 mg/kg dose when compared with the 0.25mg/kg dose (18); however, our network meta-analysis did not reaffirm this concern and instead demonstrated that all doses of tenecteplase were at least as safe as alteplase, a finding consistent with EXTEND IA TNK Part 2 where 0.25 mg/kg dose was directly compared against 0.4 mg/kg for the first time (29). The results of NOR-TEST 2 thus can be attributed to a smaller sample size and hence an inferior power of generalizability of the study.

On top of our results that provide evidence on greater efficacy of tenecteplase at 0.25 mg/kg dose, tenecteplase carries practical advantages as well. Tenecteplase owes this advantage to a lower cost, higher incidence of rehabilitation, lesser frequency of rehospitalization, faster recovery, and lower rates of dependence on palliative care in the long run (30,31). Moreover, tenecteplase may also carry a potential advantage of being efficacious at prolonged needle times. This might allow clinicians to carry out thrombolysis for up to 24 hours after stroke and salvage all viable brain tissue at longer intervals where alteplase is ineffective. This is especially beneficial for wake-up strokes i.e. strokes that occur during sleep and hence are not eligible for thrombolysis since the time since stroke onset is unknown. If found effective for 24 hours, tenecteplase will radically transform the management of wake-up strokes as well as late presentations of stroke. These properties of tenecteplase are now extensively being studied in the TWIST, TIMELESS and CHABLIS-T trials (32–34).

We believe our findings open up new avenues for clinicians and researchers to debate on and explore in this topic. To the best of our knowledge, a systematic categorization of tenecteplase into dose groups and their direct pairwise comparison with alteplase for all outcomes has not been performed before and is a defining feature of this meta-analysis. This is in addition to the fact that this meta-analysis is the largest pooling of RCT data on this topic to date. The exclusion of observational studies from our analysis has also contributed to the absence of heterogeneity from our results in large part, overcoming a limitation that has been consistently reported in previous meta-analyses (6,20), thus limiting their generalizability.

### Limitations

Limitations of this meta-analysis arise primarily from limited data, largely due to the novelty of the topic, with several important outcomes inconsistently reported across included trials. Important clinical data such as needle time was reported in varying formats that hindered any attempt to run analysis on its basis and establish a trend, if any. Moreover, an unwanted corollary of stratifying tenecteplase doses has been a decreased sample size for each of the dose tiers, thus decreasing the power of our evidence in the 0.1 mg/kg and 0.4 mg/kg tiers. However, we believe that it is a compromise worth making, owing to the fact that a common pool of data would only provide misleading results and not prove useful for clinical practice as well as research.

## Conclusions

Tenecteplase at the dose of 0.25 mg/kg is associated with higher rates of both excellent functional outcome and early neurological improvement, with a safety profile similar to alteplase. Given its superior pharmacokinetics, tenecteplase is a potentially sound alternative to alteplase. However, more trials are needed to explore questions that remain unanswered, including a direct comparison of tenecteplase with thrombectomy and the impact of NIHSS scores on the efficacy and safety profiles of tenecteplase.

## Data Availability

All data produced in the present work are contained in the manuscript.

## Author Contributions

**Aqeeb Ur Rehman:** Conceptualization, Methodology, Formal analysis, Resources, Writing - Review & Editing, Supervision.

**Aleenah Mohsin:** Formal analysis, Data curation, Writing-Original draft preparation.

**Huzaifa Ahmad Cheema:** Conceptualization, Methodology, Resources, Writing - Review & Editing, Supervision.

**Afra Zahid:** Formal analysis, Investigation, Writing-Original draft preparation.

**Muhammad Ebaad Ur Rehman:** Investigation, Writing-Original draft preparation.

**Muhammad Zain Ameer:** Validation, Writing-Original draft preparation.

**Muhammad Ayyan:** Conceptualization, Data curation, Writing-Original draft preparation.

**Muhammad Ehsan:** Methodology, Writing-Reviewing and Editing, Visualization.

**Abia Shahid:** Conceptualization, Investigation, Writing - Review & Editing.

**Muhammad Aemaz Ur Rehman:** Resources, Writing - Review & Editing, Project administration.

**Jaffer Shah:** Writing - Review & Editing, Project administration.

**Ayaz Khawaja:** Writing - Review & Editing, Supervision, Project administration.

Final approval of the version to be published: All authors.

All authors agree to be accountable for all aspects of the work.

### Statements and Declarations

### Financial support

No financial support was received for this study.

## Acknowledgements

Not applicable.

## Human and animal participants

Research involving human participants and/or animals: No animals or human subjects were used in the current study.

### Informed consents

No informed consents were required for the purpose of the current study.

### Availability of data

The data that support the findings of this study are available from the corresponding author upon reasonable request.

## References

1. Campbell BCV. Thrombolysis and Thrombectomy for Acute Ischemic Stroke: Strengths and Synergies. Semin Thromb Hemost. 2017 Mar;43(2):185–90.

2. Dhillon S. Alteplase. CNS Drugs. 2012 Oct 1;26(10):899–926.

3. Tanswell P, Modi N, Combs D, Danays T. Pharmacokinetics and Pharmacodynamics of Tenecteplase in Fibrinolytic Therapy of Acute Myocardial Infarction. Clin Pharmacokinet. 2002 Dec 1;41(15):1229–45.

4. Assessment of the Safety and Efficacy of a New Thrombolytic (ASSENT-2) Investigators, Van De Werf F, Adgey J, Ardissino D, Armstrong PW, Aylward P, et al. Single-bolus tenecteplase compared with front-loaded alteplase in acute myocardial infarction: the ASSENT-2 double-blind randomised trial. Lancet Lond Engl. 1999 Aug 28;354(9180):716–22.

5. Potla N, Ganti L. Tenecteplase vs. alteplase for acute ischemic stroke: a systematic review. Int J Emerg Med. 2022 Jan 4;15(1):1.

6. Ma P, Zhang Y, Chang L, Li X, Diao Y, Chang H, et al. Tenecteplase vs. alteplase for the treatment of patients with acute ischemic stroke: a systematic review and meta-analysis. J Neurol. 2022 Oct 1;269(10):5262–71.

7. Menon BK, Buck BH, Singh N, Deschaintre Y, Almekhlafi MA, Coutts SB, et al. Intravenous tenecteplase compared with alteplase for acute ischaemic stroke in Canada (AcT): a pragmatic, multicentre, open-label, registry-linked, randomised, controlled, non-inferiority trial. The Lancet. 2022 Jul 16;400(10347):161–9.

8. PRISMA [Internet]. [cited 2022 Feb 11]. Available from: http://prisma-statement.org/PRISMAstatement/checklist.aspx

9. The Cochrane Collaboration’s tool for assessing risk of bias in randomised trials | The BMJ [Internet]. [cited 2022 Sep 20]. Available from: https://www.bmj.com/content/343/bmj.d5928

10. Bucher HC, Guyatt GH, Griffith LE, Walter SD. The results of direct and indirect treatment comparisons in meta-analysis of randomized controlled trials. J Clin Epidemiol. 1997 Jun 1;50(6):683–91.

11. Indirect comparisons of competing interventions. Health Technol Assess [Internet]. 2005 Jul 27 [cited 2022 Sep 24];9(26). Available from: https://www.journalslibrary.nihr.ac.uk/hta/hta9260/

12. Haley EC, Thompson JLP, Grotta JC, Lyden PD, Hemmen TG, Brown DL, et al. Phase IIB/III Trial of Tenecteplase in Acute Ischemic Stroke. Stroke. 2010 Apr;41(4):707–11.

13. Alteplase versus tenecteplase for thrombolysis after ischaemic stroke (ATTEST): a phase 2, randomised, open-label, blinded endpoint study - The Lancet Neurology [Internet]. [cited 2022 Sep 24]. Available from: https://www.thelancet.com/journals/laneur/article/PIIS1474-4422(15)70017-7/fulltext

14. A Randomized Trial of Tenecteplase versus Alteplase for Acute Ischemic Stroke | NEJM [Internet]. [cited 2022 Sep 24]. Available from: https://www.nejm.org/doi/10.1056/NEJMoa1109842

15. Logallo N, Novotny V, Assmus J, Kvistad CE, Alteheld L, Rønning OM, et al. Tenecteplase versus alteplase for management of acute ischaemic stroke (NOR-TEST): a phase 3, randomised, open-label, blinded endpoint trial. Lancet Neurol. 2017 Oct;16(10):781–8.

16. Campbell BCV, Mitchell PJ, Churilov L, Yassi N, Kleinig TJ, Dowling RJ, et al. Tenecteplase versus Alteplase before Thrombectomy for Ischemic Stroke. N Engl J Med. 2018 Apr 26;378(17):1573–82.

17. Bivard A, Zhao H, Churilov L, Campbell BCV, Coote S, Yassi N, et al. Comparison of tenecteplase with alteplase for the early treatment of ischaemic stroke in the Melbourne Mobile Stroke Unit (TASTE-A): a phase 2, randomised, open-label trial. Lancet Neurol. 2022 Jun 1;21(6):520–7.

18. Kvistad CE, Næss H, Helleberg BH, Idicula T, Hagberg G, Nordby LM, et al. Tenecteplase versus alteplase for the management of acute ischaemic stroke in Norway (NOR-TEST 2, part A): a phase 3, randomised, open-label, blinded endpoint, non-inferiority trial. Lancet Neurol. 2022 Jun 1;21(6):511–9.

19. Li S, Pan Y, Wang Z, Liang Z, Chen H, Wang D, et al. Safety and efficacy of tenecteplase versus alteplase in patients with acute ischaemic stroke (TRACE): a multicentre, randomised, open label, blinded-endpoint (PROBE) controlled phase II study. Stroke Vasc Neurol [Internet]. 2022 Feb 1 [cited 2022 Sep 24];7(1). Available from: https://svn.bmj.com/content/7/1/47

20. Oliveira M, Fidalgo M, Fontão L, Antão J, Marques S, Afreixo V, et al. Tenecteplase for thrombolysis in stroke patients: Systematic review with meta-analysis. Am J Emerg Med. 2021 Apr;42:31–7.

21. Demchuk AM, Tanne D, Hill MD, Kasner SE, Hanson S, Grond M, et al. Predictors of good outcome after intravenous tPA for acute ischemic stroke. Neurology. 2001 Aug 14;57(3):474–80.

22. Emberson J, Lees KR, Lyden P, Blackwell L, Albers G, Bluhmki E, et al. Effect of treatment delay, age, and stroke severity on the effects of intravenous thrombolysis with alteplase for acute ischaemic stroke: a meta-analysis of individual patient data from randomised trials. Lancet Lond Engl. 2014 Nov 29;384(9958):1929–35.

23. Kheiri B, Osman M, Abdalla A, Haykal T, Ahmed S, Hassan M, et al. Tenecteplase versus alteplase for management of acute ischemic stroke: a pairwise and network meta-analysis of randomized clinical trials. J Thromb Thrombolysis. 2018 Nov;46(4):440–50.

24. Powers WJ, Derdeyn CP, Biller J, Coffey CS, Hoh BL, Jauch EC, et al. 2015 American Heart Association/American Stroke Association Focused Update of the 2013 Guidelines for the Early Management of Patients With Acute Ischemic Stroke Regarding Endovascular Treatment. Stroke. 2015 Oct;46(10):3020– 35.

25. Mistry EA, Mistry AM, Nakawah MO, Chitale RV, James RF, Volpi JJ, et al. Mechanical Thrombectomy Outcomes With and Without Intravenous Thrombolysis in Stroke Patients. Stroke. 2017 Sep;48(9):2450–6.

26. Wu X, Ge Y, Chen S, Yan Z, Wang Z, Zhang W, et al. Thrombectomy with or without thrombolysis in patients with acute ischemic stroke: a systematic review and meta-analysis. J Neurol. 2022 Apr;269(4):1809–16.

27. Could mechanical thrombectomy replace thrombolysis in the treatment of acute and subacute limb ischemia? - Minerva Cardioangiologica 2019 June;67(3):234-45 [Internet]. [cited 2022 Sep 20]. Available from: https://www.minervamedica.it/en/journals/minerva-cardiology-angiology/article.php?cod=R05Y2019N03A0234

28. Campbell BCV, Kappelhof M, Fischer U. Role of Intravenous Thrombolytics Prior to Endovascular Thrombectomy. Stroke. 2022 Jun;53(6):2085–92.

29. Campbell BCV, Mitchell PJ, Churilov L, Yassi N, Kleinig TJ, Dowling RJ, et al. Effect of Intravenous Tenecteplase Dose on Cerebral Reperfusion Before Thrombectomy in Patients With Large Vessel Occlusion Ischemic Stroke: The EXTEND-IA TNK Part 2 Randomized Clinical Trial. JAMA. 2020 Apr 7;323(13):1257–65.

30. Gao L, Moodie M, Mitchell PJ, Churilov L, Kleinig TJ, Yassi N, et al. Cost-Effectiveness of Tenecteplase Before Thrombectomy for Ischemic Stroke. Stroke. 2020 Dec;51(12):3681–9.

31. Nepal G, Kharel G, Ahamad ST, Basnet B. Tenecteplase versus Alteplase for the Management of Acute Ischemic Stroke in a Low-income Country-Nepal: Cost, Efficacy, and Safety. Cureus. 2018 Feb 9;10(2):e2178.

32. Roaldsen MB, Lindekleiv H, Eltoft A, Jusufovic M, Søyland MH, Petersson J, et al. Tenecteplase in wake-up ischemic stroke trial: Protocol for a randomized-controlled trial. Int J Stroke. 2021 Oct 1;16(8):990–4.

33. Genentech, Inc. A Phase III, Prospective, Double-blind, Randomized, Placebo-controlled Trial of Thrombolysis in Imaging-eligible, Late-window Patients to Assess the Efficacy and Safety of Tenecteplase (TIMELESS) [Internet]. clinicaltrials.gov; 2022 Sep [cited 2022 Sep 22]. Report No.: results/NCT03785678. Available from: https://clinicaltrials.gov/ct2/show/results/NCT03785678

34. Dong Q. Prospective, Multicenter, Open, End-point Blinded, Stratified Block Randomized, Parallel Positive Controlled Clinical Trial of Tenecteplase in Acute Ischemic Stroke With Large Vessel Occlusion Over Time Window [Internet]. clinicaltrials.gov; 2022 Sep [cited 2022 Sep 22]. Report No.: NCT04516993. Available from: https://clinicaltrials.gov/ct2/show/NCT04516993

