## Supplementary Table 1 for "Comparative Efficacy and Safety of Tenecteplase and Alteplase in Acute Ischemic Stroke: A Pairwise and Network Meta-analysis of Randomized Controlled Trials"

**Detailed Search Strategy**

**Supplementary Table 1.** Risk of Bias Assessment of included RCTs.

**Supplementary Figure 1.** Forest plot for good functional outcome at 3 months (mRS 0-2).

**Supplementary Figure 2.** Meta regression plot of the effect of baseline NIHSS score on excellent functional outcome (mRS 0-1) for all doses combined.

**Supplementary Figure 3.** Meta regression plot of the effect of baseline median NIHSS score on good functional outcome (mRS 0-2) for all doses combined.

**Supplementary Figure 4.** Meta regression plot of the effect of baseline NIHSS score on excellent functional outcome (mRS 0-1) at 0.25 mg/kg dose.

**Supplementary Figure 5.** Meta regression plot of the effect of baseline median NIHSS score on good functional outcome (mRS 0-2) at 0.25 mg/kg dose.

**Supplementary Figure 6.** Forest plot for mortality at 3 months.

**Supplementary Figure 7.** Meta regression plot of the effect of baseline median NIHSS score on mortality at 3 months for all doses combined.

**Supplementary Figure 8.** Meta regression plot of the effect of baseline median NIHSS score on mortality at 3 months at 0.25 mg/kg dose.

**Supplementary Figure 9.** Forest plot for intracerebral hemorrhage (ICH)

**Supplementary Figure 10.** Forest plot for symptomatic Intracerebral hemorrhage (sICH)

**Supplementary Figure 11.** Network meta-analysis on excellent functional outcome (mRS 0-1)

**Supplementary Figure 12.** Network meta-analysis on good functional outcome (mRS 0-2)

**Supplementary Figure 13.** Network meta-analysis on symptomatic intracerebral hemorrhage (sICH)

**Supplementary Figure 14.** Network meta-analysis on mortality at 3 months

**Detailed Search Strategy**

**Search strategy for PubMed**

1. Tenecteplase (Mesh)
2. Tenecteplase (tiab)
3. Tenecteplase (ot)
4. TNK (tiab)
5. TNKase (tiab)
6. TNK-tPA (tiab)
7. Metylase (tiab)
8. Recombinant human TNK tissue-type plasminogen activator (tiab)
9. rhTNK-Tpa (tiab)
10. 1 OR 2 OR 3 OR 4 OR 5 OR 6 OR 7 0R 8 OR 9
11. Alteplase (Mesh)
12. Alteplase (tiab)
13. Tissue Plasminogen Activator (Mesh)
14. Tissue Plasminogen Activator (tiab)
15. Plasminogen activator (tiab)
16. T plasminogen activator (tiab)
17. Tissue type plasminogen activator (tiab)
18. R-tPA (tiab)
19. Activase (tiab)
20. 11 OR 12 OR 13 OR 14 OR 15 OR 16 OR 17 OR 18 OR 19
21. Stroke (Mesh)
22. Stroke (tiab)
23. Acute ischemic stroke (Mesh)
24. Acute ischemic stroke (tiab)
25. Ischemic stroke (tiab)
26. Acute stroke (tiab)
27. Cerebrovascular accident (tiab)
28. Cerebrovascular event (tiab)
29. Brain vascular accident (tiab)
30. Brain vascular event (tiab)
31. 21 OR 22 OR 23 OR 24 OR 25 OR 26 OR 27 OR 28 OR 29 OR 30
32. (10 AND 20 AND 31) NOT (animals)

**Search strategy for Embase**

1. tenecteplase: ti.ab
2. tenecteplase/exp
3. TNK: ti.ab
4. TNKase: ti.ab
5. TNK-tPA: ti.ab
6. metylase: ti.ab
7. RhTNK-tPA: ti.ab
8. 1 OR 2 OR 3 OR 4 OR 5 OR 6 OR 7
9. alteplase/exp
10. alteplase: ti.ab
11. r-tPA: ti.ab
12. tissue plasminogen activator: ti.ab
13. activase: ti.ab
14. 9 OR 10 OR 11 OR 12 OR 13
15. stroke/exp
16. stroke: ti.ab
17. acute ischemic stroke: ti.ab
18. acute stroke: ti.ab
19. ischemic stroke: ti.ab
20. 15 OR 16 OR 17 OR 18 OR 19
21. 8 AND 14 AND 20

**Search strategy for Cochrane**

1. MeSH descriptor: Tenecteplase
2. Tenecteplase: ti.ab.kw
3. TNK: ti.ab.kw
4. TNKase: ti.ab.kw
5. TNK-tPA: ti.ab.kw
6. RhTNK-tPA: ti.ab.kw
7. Metylase: ti.ab.kw
8. 1 OR 2 OR 3 OR 4 OR 5 OR 6 OR 7
9. MeSH descriptor: Alteplase
10. Alteplase: ti.ab.kw
11. r-tPA: ti.ab.kw
12. Tissue plasminogen activator: ti.ab.kw
13. Plasminogen activator: ti.ab.kw
14. Acticase: ti.ab.kw
15. 9 OR 10 OR 11 OR 12 OR 13 OR 14
16. MeSH descriptor: Stroke
17. Stroke: ti.ab.kw
18. Acute ischemic stroke: ti.ab.kw
19. Ischemic stroke: ti.ab.kw
20. Acute stroke: ti.ab.kw
21. 16 OR 17 OR 18 OR 19 OR 20
22. 8 AND 15
23. AND 21

**Supplementary Table 1. Risk of Bias Assessment of included RCTs**

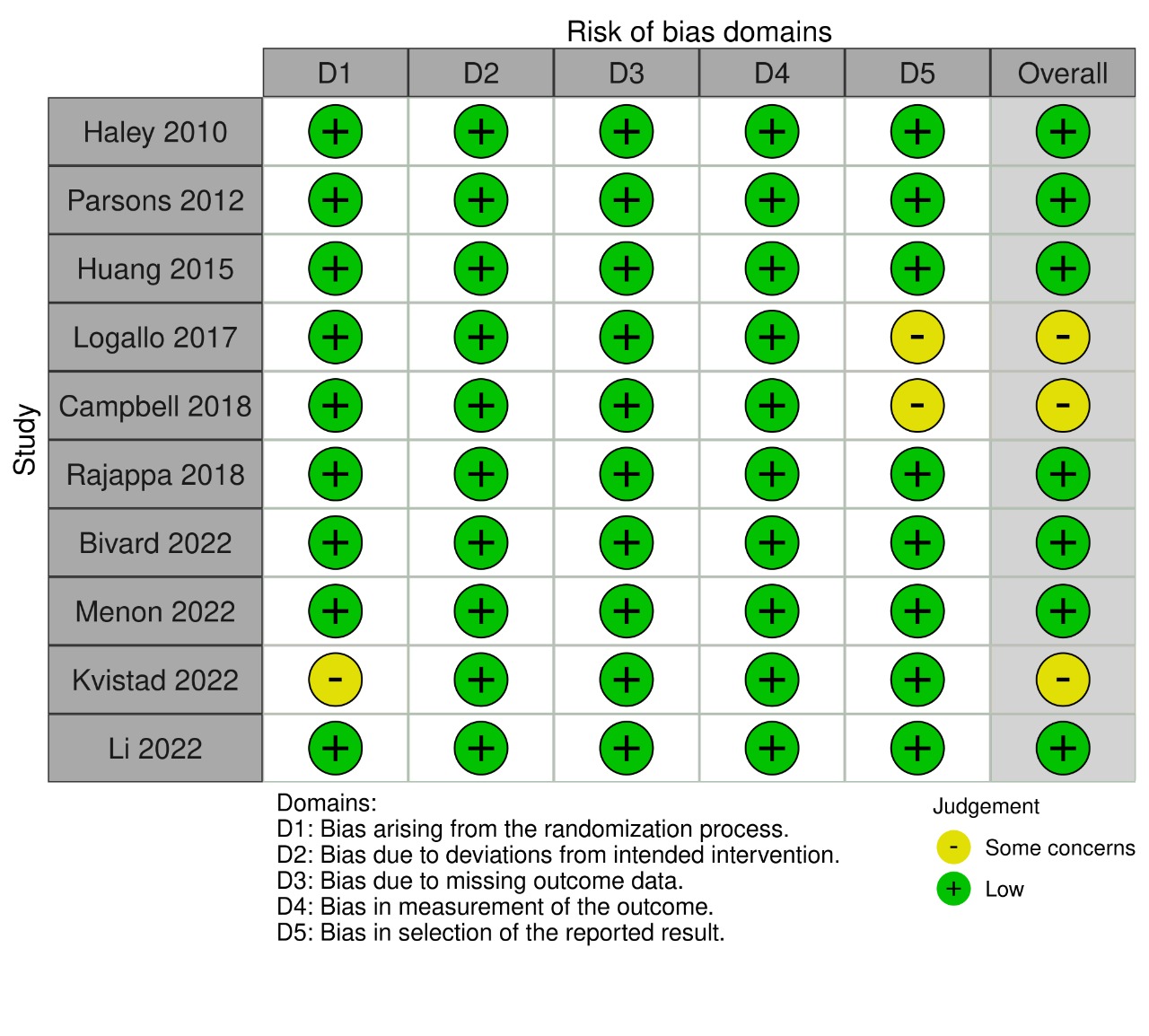

**Supplementary Figure 1. Forest plot for good functional outcome at 3 months (mRS 0-2)**

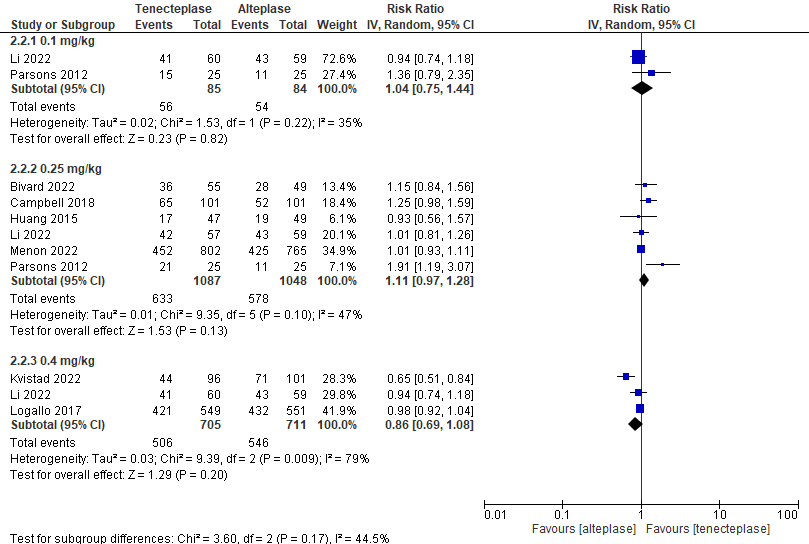

**Supplementary Figure 2. Meta regression plot of the effect of baseline median NIHSS score on excellent functional outcome (mRS 0-1)**

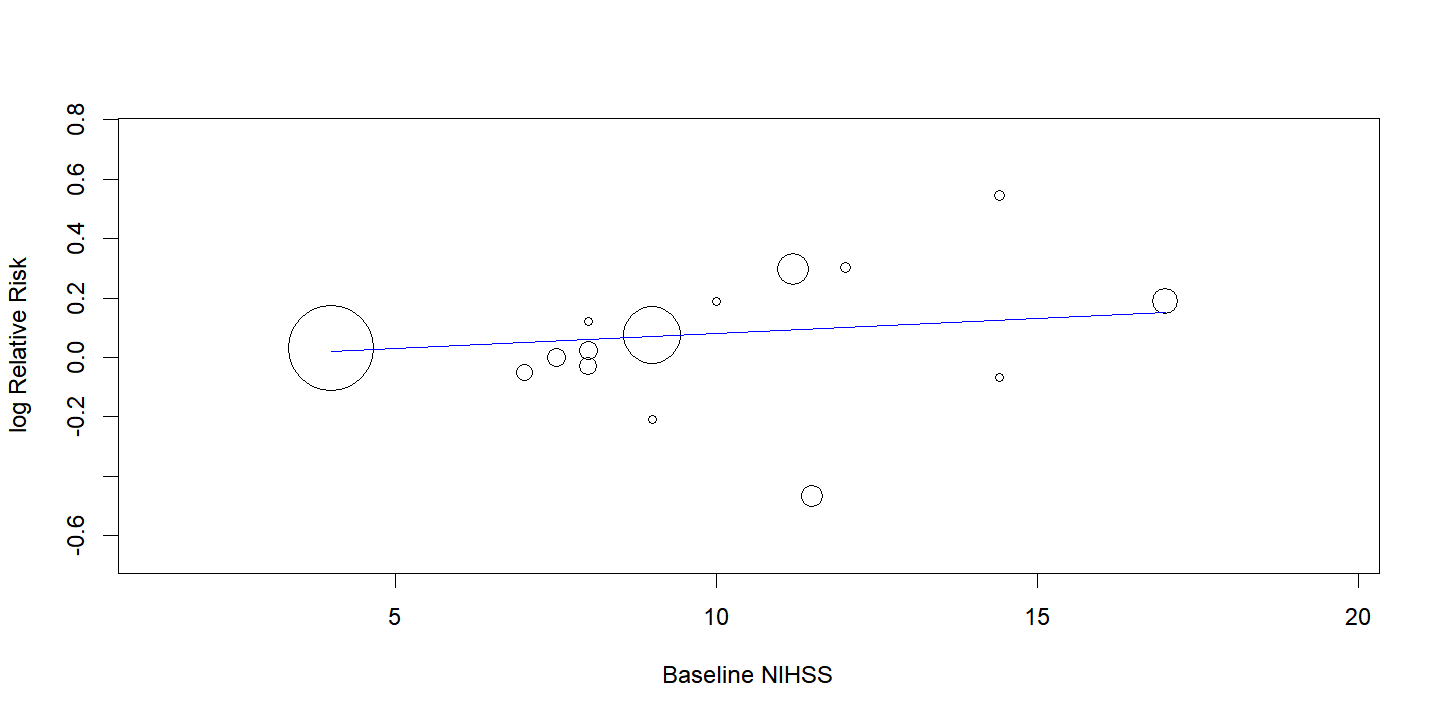

**p-value= 0.258**

**Supplementary Figure 3. Meta regression plot of the effect of baseline median NIHSS score on good functional outcome (mRS 0-2)**

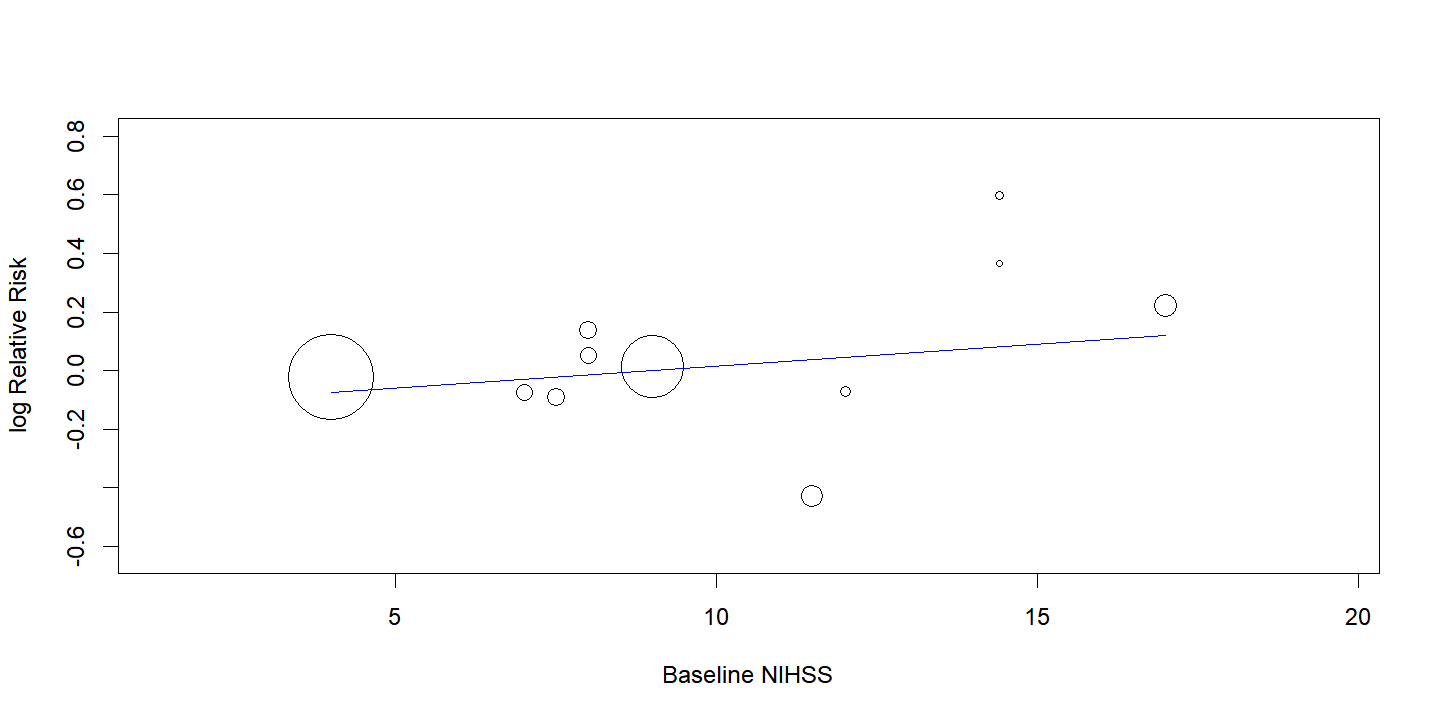

**p-value= 0.271**

**Supplementary Figure 4. Meta regression plot of the effect of baseline median NIHSS score on excellent functional outcome (mRS 0-1) at 0.25 mg/kg dose**

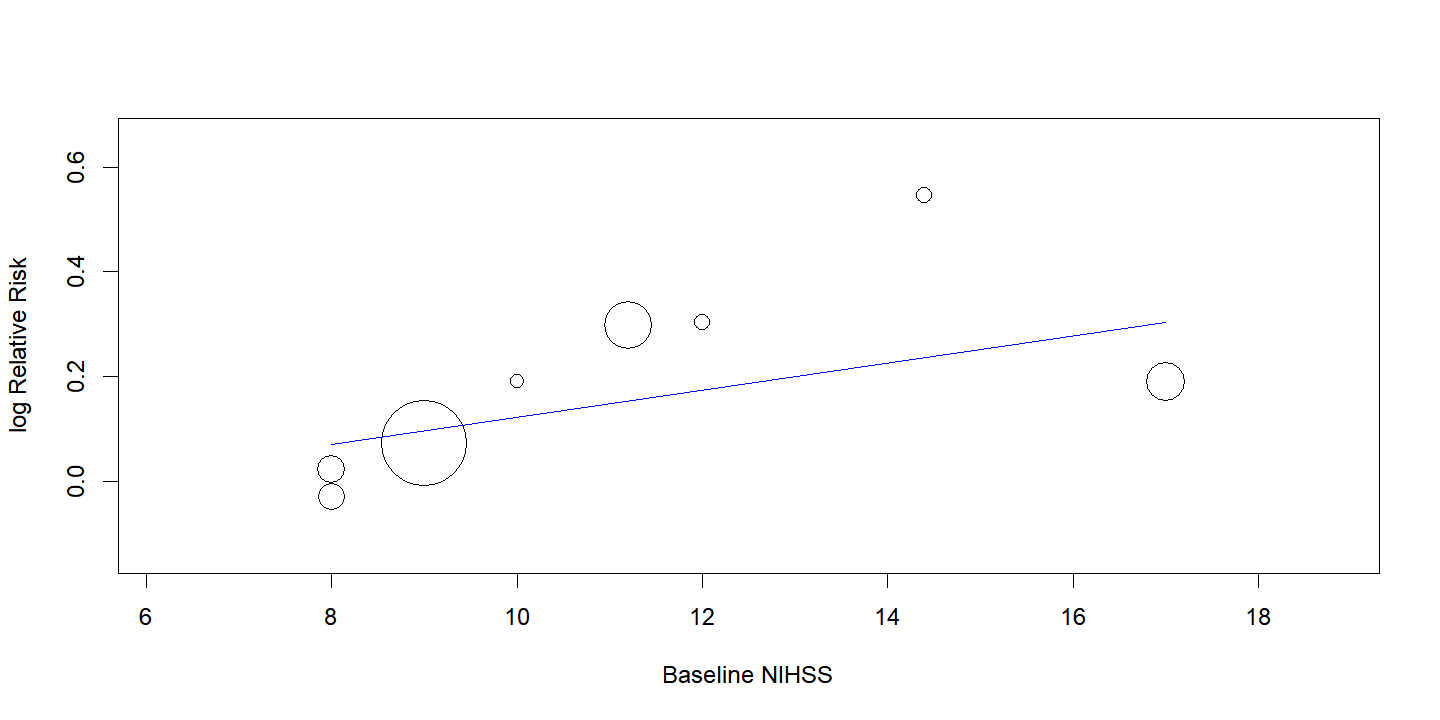

**p-value=0.171**

**Supplementary Figure 5. Meta regression plot of the effect of baseline median NIHSS score on good functional outcome (mRS 0-2) at 0.25 mg/kg dose**
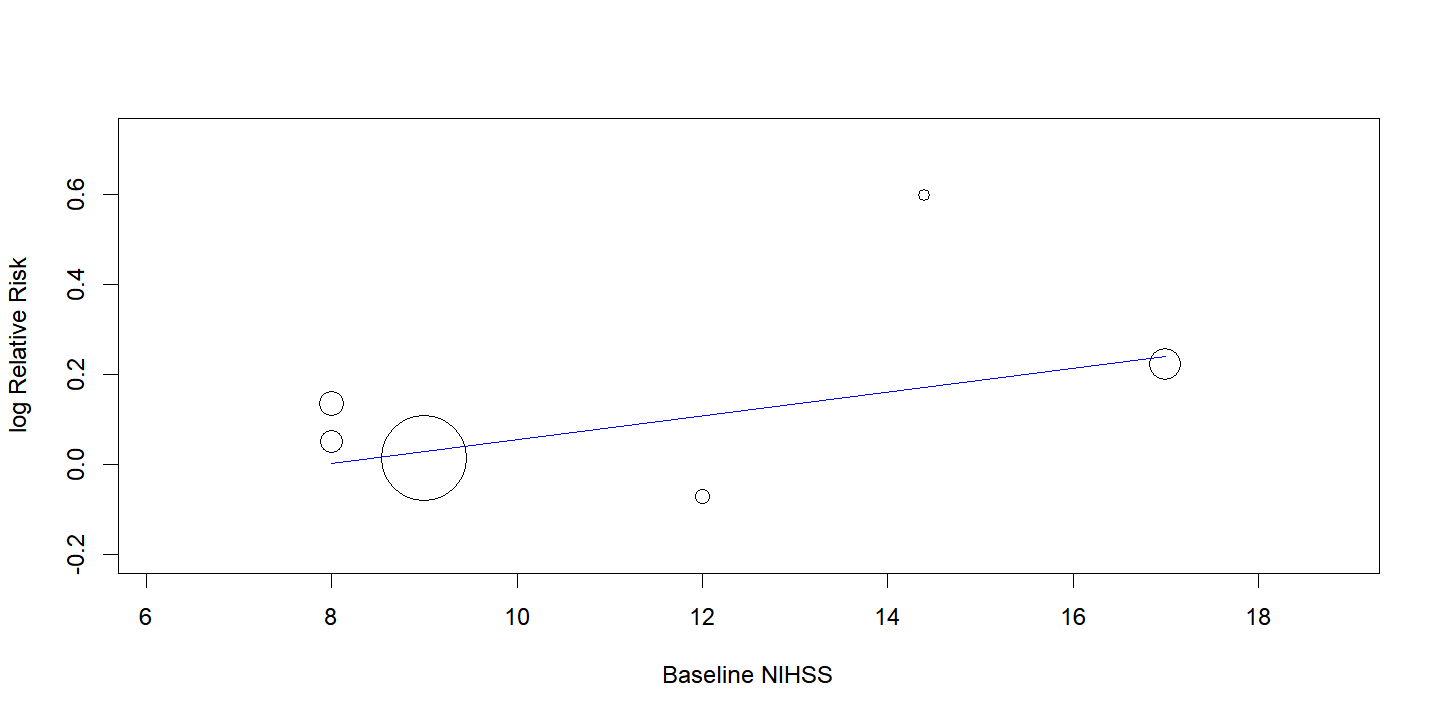

**p-value= 0.084**

**Supplementary Figure 6. Forest plot for mortality at 3 months**

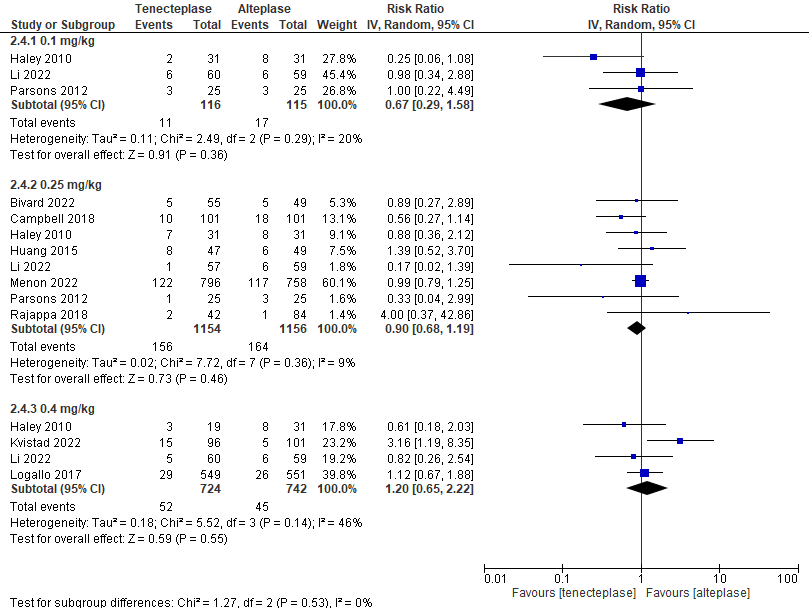

**Supplementary Figure 7. Meta regression plot of the effect of baseline median NIHSS score on mortality at 3 months**

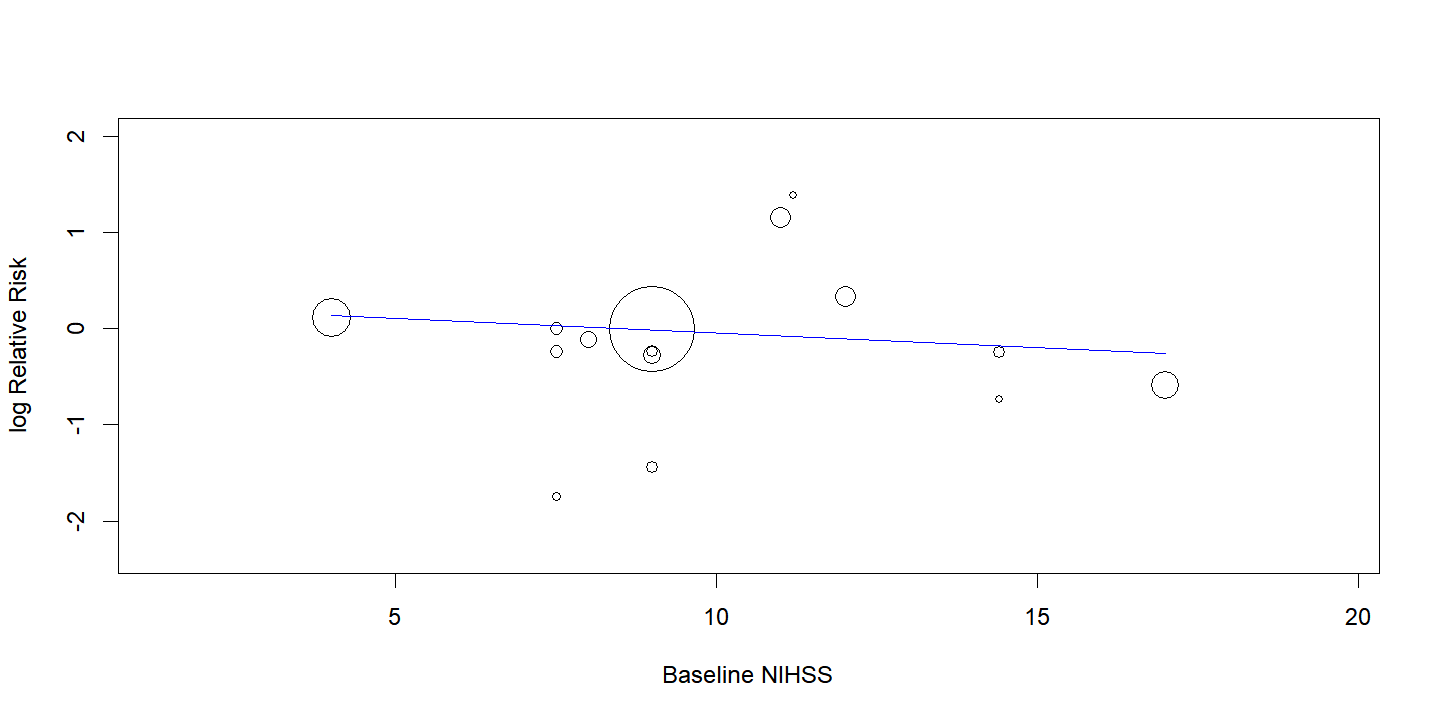

**p-value= 0.356**

**Supplementary Figure 8. Meta regression plot of the effect of baseline median NIHSS score on mortality at 3 months at 0.25 mg/kg dose**

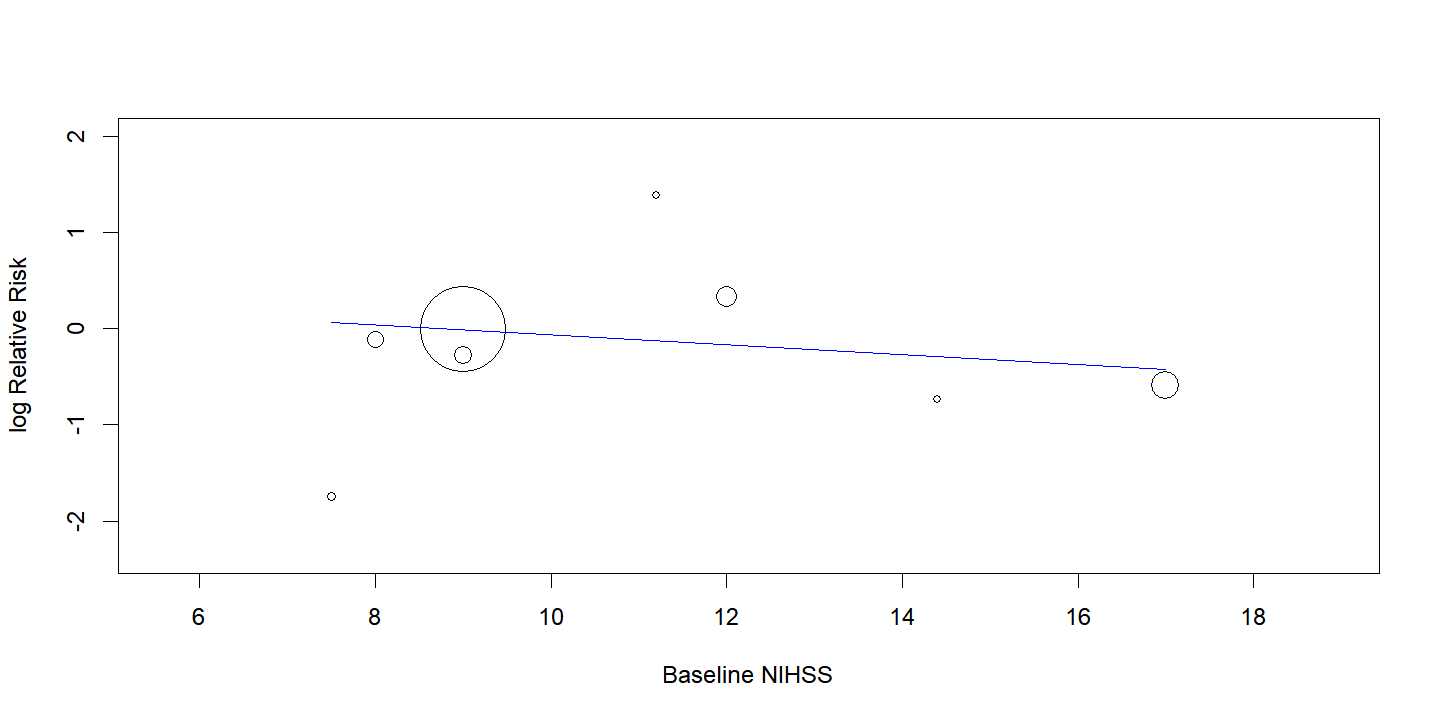

**p-value= 0.264**

**Supplementary Figure 9. Forest plot for Intracerebral hemorrhage (ICH)**

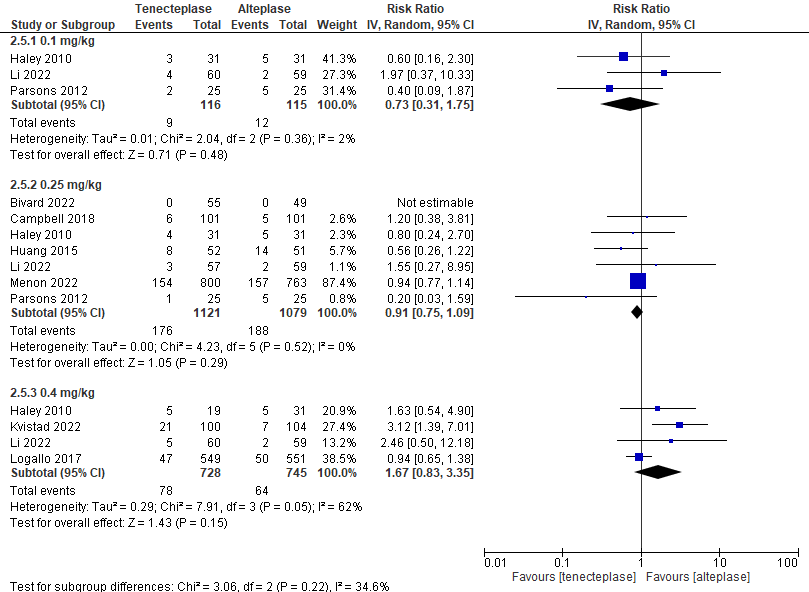

**Supplementary Figure 10. Forest plot for symptomatic Intracerebral hemorrhage (sICH)**

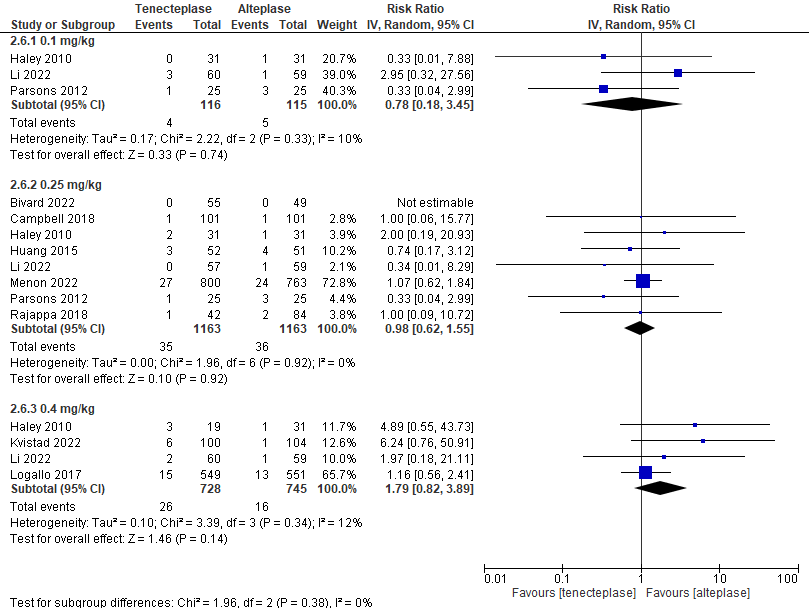

**Supplementary Figure 11. Network meta-analysis on excellent functional outcome (mRS 0-1)**

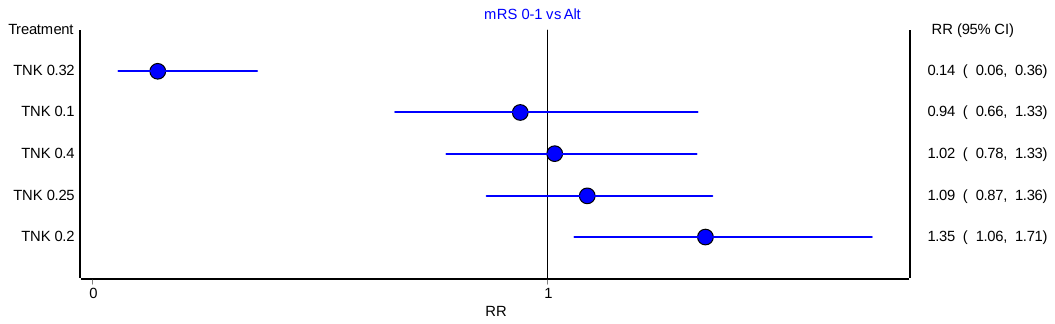

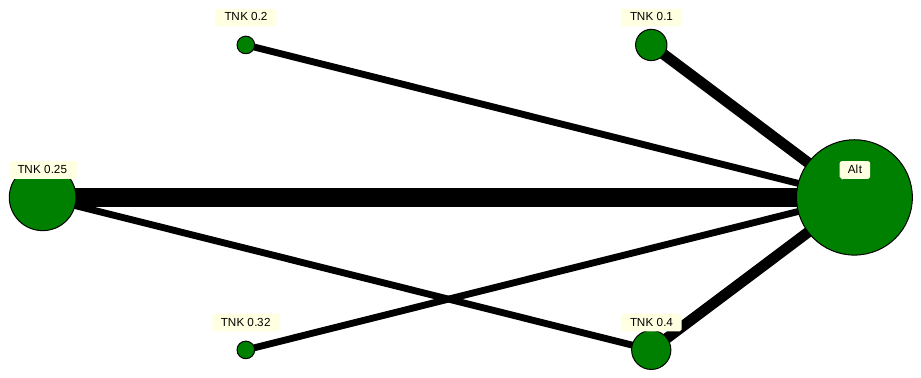

**Supplementary Figure 12. Network meta-analysis on good functional outcome (mRS 0-2)**

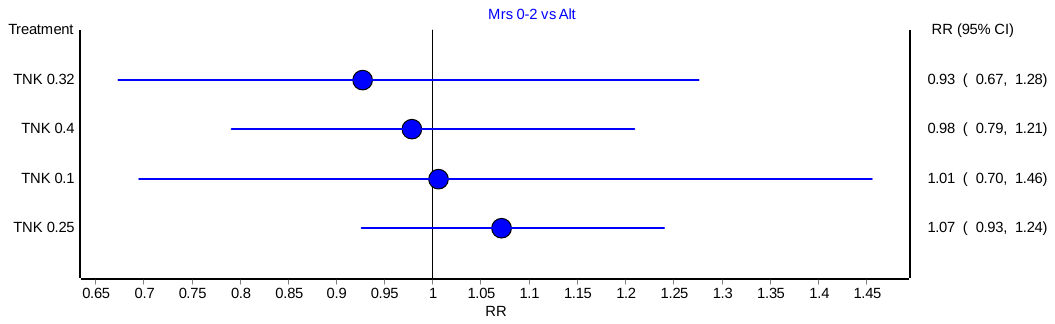

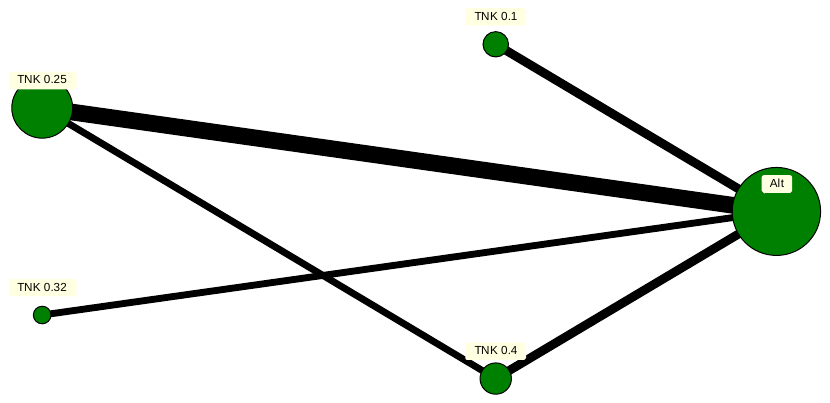

**Supplementary Figure 13. Network meta-analysis on symptomatic intracerebral hemorrhage (sICH)**

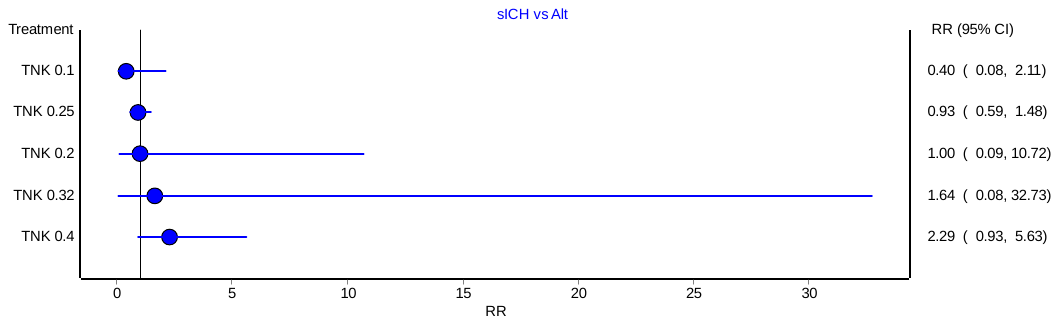

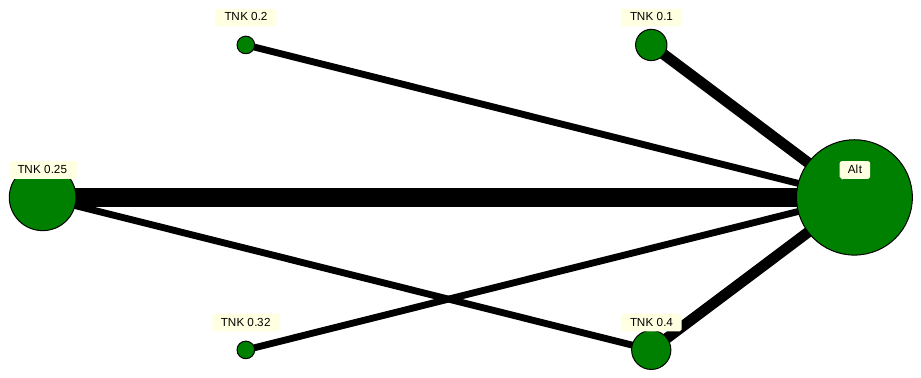

**Supplementary Figure 14. Network meta-analysis on mortality**

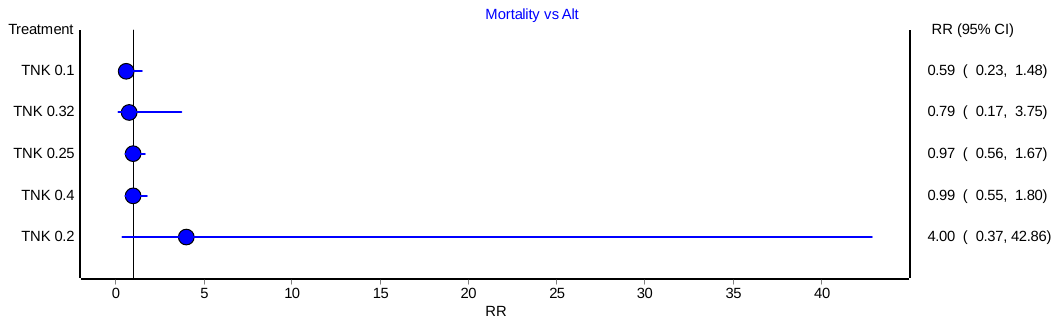

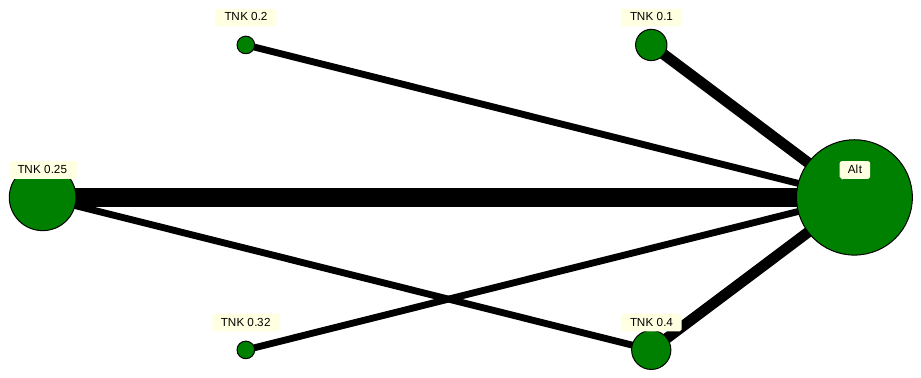
